# Understanding Waiting Lists Pressures

**DOI:** 10.1101/2022.08.23.22279117

**Authors:** Kevin Fong, Yasser Mushtaq, Thomas House, Dan Gordon, Yingrae Chen, Darren Griffths, Shazaad Ahmad, Neil Walton

## Abstract

NHS waiting lists currently sit at record lengths due to a combination of the immediate impact of the pandemic and, as well as, long-run pressures requiring investment on NHS resources. These factors have left managers and clinicians with increasingly complex decisions when scheduling elective operations. It is imperative that managers understand the basic dynamics, tradeoffs, and pressures when managing waiting lists.

Queueing theory is a key part of operational research, extensively used throughout manufacturing, retail, information technology and other sectors. This article provides an exposition of the theory of queues within the context of the current NHS backlog. With this information a manager will be able understand the demand, queue size, waiting times, capacity requirements and trade-offs for different waiting lists. We describe the metrics and a reporting system developed to understand waiting list pressures in a large NHS trust. Our aim is to enable managers to better understand their waiting lists, to achieve targets and improve health outcomes.

---

It has arguably never been more important for clinicians and NHS managers to understand and manage their waiting lists. Here, we seek to facilitate this process by describing several mathematical principles that govern the behaviour of queues. These are drawn from well-established scientific approaches to scheduling and reflect the real-life challenges that NHS Trusts face. Importantly, such principles imply certain hard limits on performance. Some results are intuitively clear while others can at first be counter-intuitive. Nevertheless we present a robust set of metrics and measures that an NHS manager can use to understand their waiting lists, assess targets, plan capacity and route patients between different hospitals.

Queues arise whenever there is a mismatch between the demand and capacity for service. In this sense, waiting lists are queues. If the rate at which items are added outstrips the rate at which they are removed, a queue will grow indefinitely. This much is intuitively obvious. Clearly this is a situation to be avoided; however, even at a constant queue length one might ask, when is a waiting list too big then how much bigger should the rate of removals be to meet targets? For a manager, we can introduce the concept of a Target Equilibrium. A *Target Equilibrium* can be said to be achieved if the rate of addition and removal of items is appropriately matched, such that the queue is stable <u>and</u> acceptable target waiting times can be maintained.

A range of mathematical relationships govern the behaviour of queues operating within these parameters. The mathematical underpinnings are themselves somewhat complex, to the extent that Queueing Theory is itself a separate branch of mathematics. However, even the basic principles yield useful insight for managers and clinicians alike. Importantly, they can be used to determine the increase in capacity that is required to achieve targets. This helps managers to understand the limits of performance of their systems and set broad expectations regarding the timescales required to achieve operational targets.

There are several important observations arising from the results discussed below. For instance, one of the counterintuitive consequences of queuing theory is that for stable lists operating at or around target equilibrium, the list utilisation must be less than 100%. This seems to run contrary to the idea of maximising efficiency; indeed, managers are often penalised for not achieving close to 100% utilisation. But for a list to remain stable, with short waiting time targets, Queuing Theory dictates that there must be an appreciable time period where the resource is effectively fallow.

This is important: Queueing Theory defines a set of laws which cannot be violated, as they describe fundamental limits which cannot be outperformed without substantial alteration of the underlying system. And while mathematically described queues represent simplified models of real-world situations, results that we introduce, such as Little’s Law, apply to the dynamics of the waiting list in the same way that the Newton’s law of motion applies to a stone hurled into the air.

The behaviour of queues when they are close to equilibrium - i.e. operating in and around the rate of addition and removal of items from the queue is well matched allowing target waiting times to be met - is very different from those operating far from equilibrium i.e. in which demand is growing and/or the queue size is extremely long.

In the former (stable) scenario small adjustments to capacity can successfully stabilise the queue and allow acceptable waiting times to be met. In the latter, i.e. in a queue operating far from target equilibrium, extraordinary processes are required to return the queue to stability and thereby achieve acceptable waiting times in a reasonable timeframe. Here we provide tools so that a manager or clinician is aware of the symptoms of an unhealthy waiting list and so interventions are introduced well before targets are being missed.

Further, managers must make trade-offs between waiting lists. Often it may not be clear to a manager which patient, procedure, speciality, site or priority group is most at risk of missing clinical targets. We introduce a notation of *waiting list pressure* as a mechanism of comparing and understanding the pressures on different services. From this, simple routing and scheduling decisions can be conducted by a manager to equilibrate queues and meet targets.

We provide examples and descriptions of key queueing metrics and how these are applied in reporting mechanisms that are used to understand the current resource requirements and waiting list pressures across the Manchester region.

## Basics of Queueing

We describe the basic components of a queue before we proceed to explain how these should be measured and adjusted to improve efficiency. Within the context of a waiting list, the *queue size* is the number of patients waiting for their procedure. The *waiting time* of a patient is the time from referral until the operation or medical procedure is conducted. The *demand* is the number patients referred per week for a procedure. The *capacity* is the number of patients that can be served from the waiting list each week. The capacity can be measured as the number of waiting list removals each week given that the waiting list did not empty.

Another way to think of the rate of service at a queue is in terms of its load, which is the number of arrivals that occur for every patient leaving the queue. The *load* is the ratio between demand and capacity:

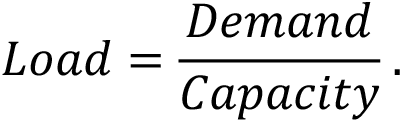

Each waiting list has a priority within the NHS. These are typically labelled P1, P2, P3 and P4, which each respectively has a target waiting time of 1-2 days, 4 weeks, 12 weeks and 52 weeks, (1).

Now that we have a some terminology in place, we describe an example which we will use to help explain our analysis.

## A Running Example

We will consider the following example throughout the text, which is based on the P4 waiting list for Ear, Nose and Throat (ENT) in the University of Manchester NHS Foundation Trust. Here there is a queue size of 1200 patients. The average waiting time currently in the queue is 63 weeks, and approximately 51% of the waiting list have already missed their target of 52 weeks. The demand is 30 patients per week and the capacity currently allocated to patients is 27 procedures per week. The standard deviation in the number of operations per week is 160. The first observation from this is that the demand is above the capacity allocated to these patients.

## Capacity, Demand and Load

In our example, the 30 patients added to the waiting list each week and 27 removed, results in a net increase of 3 patients per week. The sustained mismatch between service capacity and demand is what has led to over 1200 patients waiting for operations.

**Fact 1:** The key observation is that we require that the capacity is bigger than the demand otherwise the waiting list will grow indefinitely. So, we must have *capacity > Demand*.

Shortly we will discuss how much bigger the capacity needs to be. However, for now, Figure 1 gives a simulation of what happens to this waiting list over the next 26 weeks for ENT P4 when the waiting list’s capacity is less than the demand.

**Figure 1.**
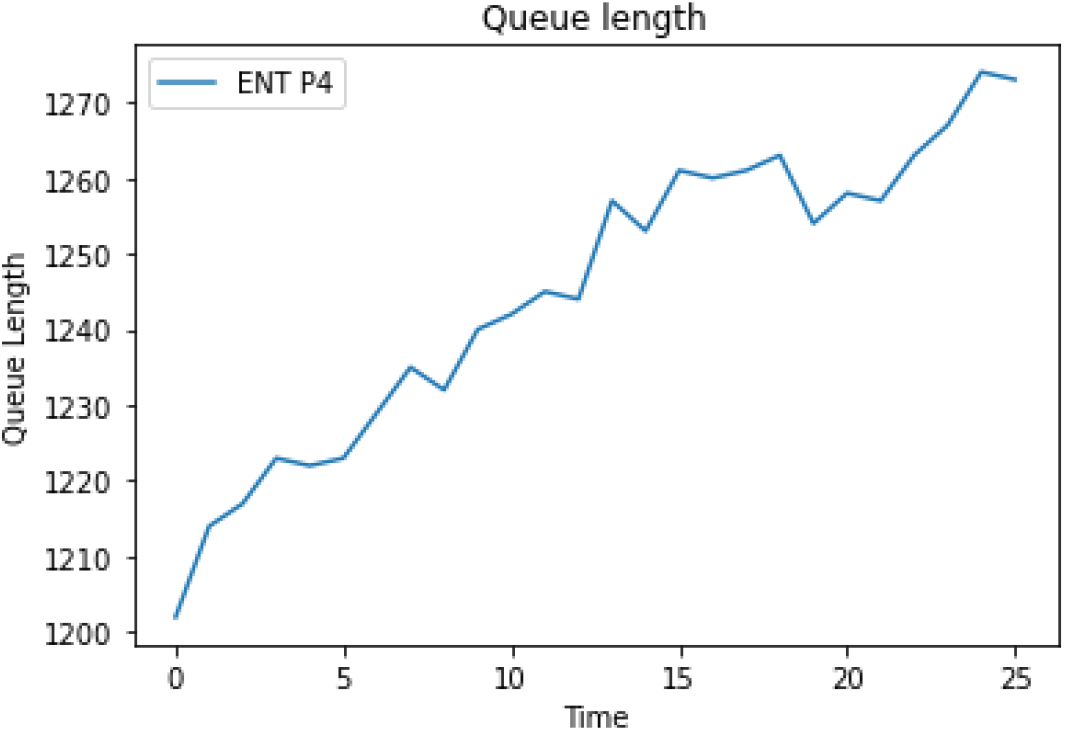
Without any intervention, the waiting list for ENT P4 grows indefinitely.

We now look at the load on the queue. For our example the load is 1.11 which is greater than 1. We see from Fact 1 that if the load is bigger than 1 then the queue becomes unstable. However, if the load is less than 1 then the queue will drain and will spend some proportion of the time empty.

Another way to think about the load, when it is less than 1, is the amount of time that work is being done on the waiting list. Assuming we always allocated capacity when there is work to be done, then this is the proportion of time that the waiting list is non-empty. To see why this is true, note that the demand is the number of referrals per week and the capacity is the rate at which the waiting list is drained, when the waiting list is not empty. If we can drain the queue and we account for the time that the waiting list is empty, the capacity allocated to the non-empty queue must balance out with the demand. This gives that

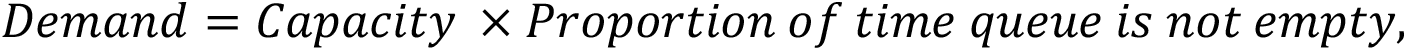

which then implies

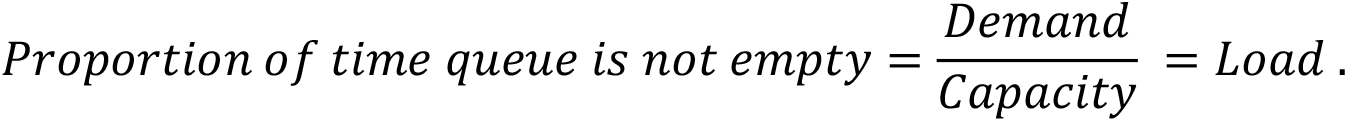

Thus, we see that for any waiting list to be stable, there must be some non-negligible proportion of time that work is *not* being done on the list (because there is no work to be done). This observation is summarized below:

**Fact 2:** If the load is greater than 1, then the queue is unstable, and the waiting list will grow indefinitely. If the load is less than 1, then the queue will be stable and the load is the proportion of the time that the waiting list is non-empty. That is when the load is less than 1

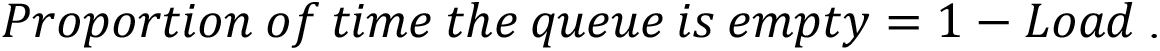

The above states that the load is essentially the utilization of the waiting list, i.e. the proportion of time that work is being done on the waiting list. As such, the above fact highlights a dichotomy between waiting times and utilization: you cannot have full utilization and low waiting times. **If you want to have low waiting times, then there must be a non-negligible fraction of time where services are *not* being used**. The mathematical implications run further such that, for lists where the target waiting time is both short and the consequences of not achieving that target are severe – for example priority 1 cancer lists, where missing targets may lead to loss of control of the disease – the list should be designed such that there is significant un-utilised time. As a rule of thumb, the shorter and more critical the target waiting time the lower the list utilisation must be to ensure that the target is consistently met.

This principle is particularly important for emergency response services – for example the ambulance service – where assets designed to attend some calls within a specified short time will require job cycle dynamics with very low utilisation. For instance, it is standard in New Zealand countries for ambulances to attain 60% utilization (2). It is recommended that bed occupancy levels are below 85% (3). This challenges a fallacy that all services must be run at 100% utilization. Indeed, for specialist services or services requiring fast response times, this could be harmful for waiting lists with performance targets.

It is important for managers to understand that this is not inefficiency, rather it is a mathematical limitation imposed by queuing theory, which requires a trade to be made, between list utilisation and instability of the queue.

## Waiting List Targets

A simple question is, how long should the typical patient wait for their operation in order for the waiting list to have a high chance of making targets? It turns out that this may be smaller than one might initially expect.

We know that the waiting list will grow in roughly a straight line if demand is constantly above the capacity allocated to the queue (recall Figure 1). Conversely, if the capacity is consistently above the mean demand for the waiting list, the waiting list will empty and the queue will reach some equilibrium behavior. Patients may still need to wait. Here the waiting time will depend on the random mismatch between the demand and capacity allocated to the waiting list. In equilibrium the waiting time is random, and, given this, it is natural to ask about the probability distribution of the waiting time of patients.

During the pandemic there was much talk of exponential *growth*. For example, if coronavirus is unchecked, then R0 is greater than 1 and the prevalence of infection doubles every 3 days. When we look at the waiting time of a queue, a related notion of exponential decay, occurs, and the load plays an analogous role to the R0 growth factor.

**Fact 3**. If the load on a queue is less than 1 then the chance of missing the target halves each time we increase the target by some fixed number of days.

The resulting probability distribution for the waiting time is called the exponential distribution. The waiting time of an underloaded queue is well modelled by an exponential distribution under a broad class of assumptions (4) (5). See Figure 2 for the exponential decay in the distribution of the waiting time.

**Figure 2.**
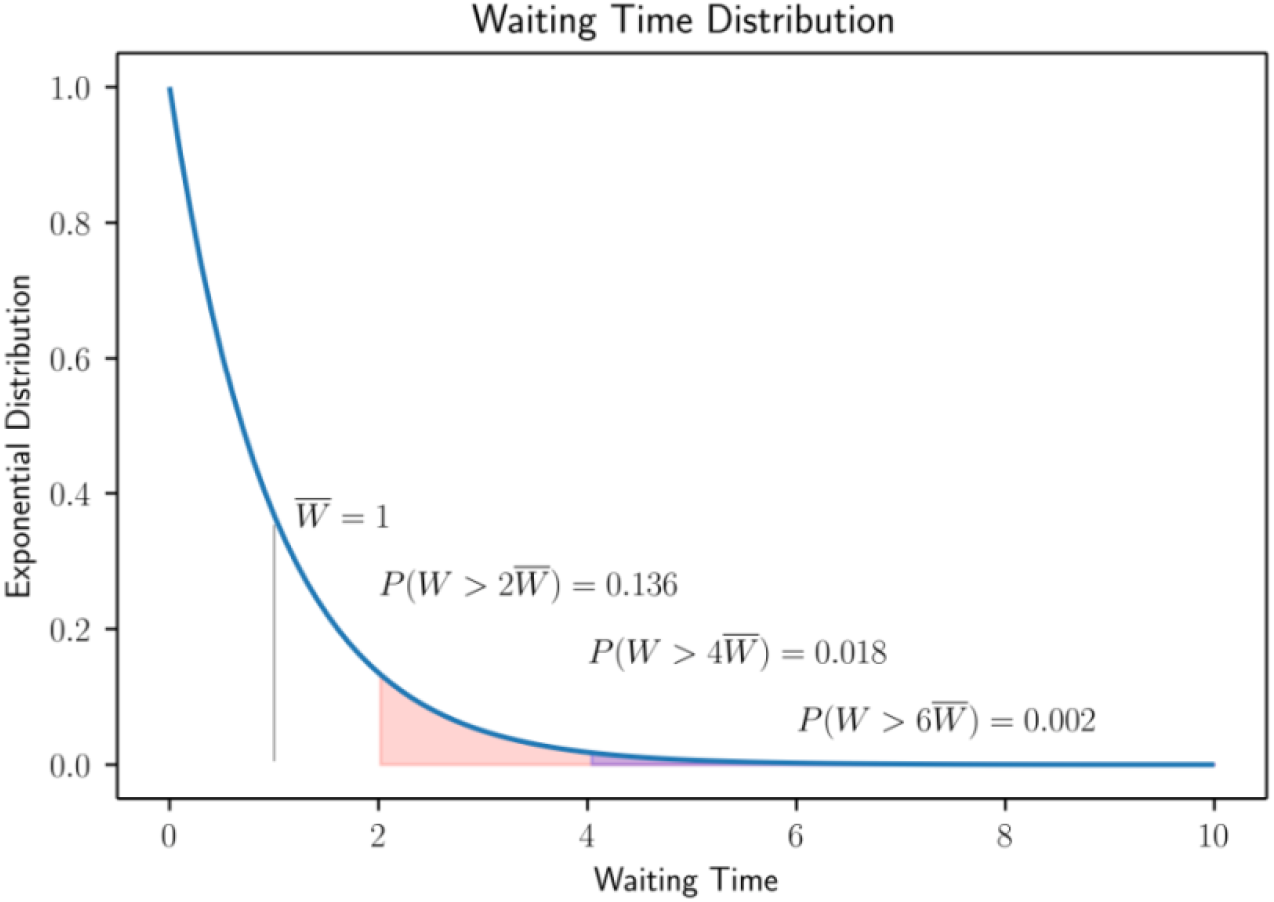
The probability distribution of waiting times for a stable queue. For a stable queue, the probability of waiting a given multiple above the mean decays exponentially fast.

**Figure 3.**
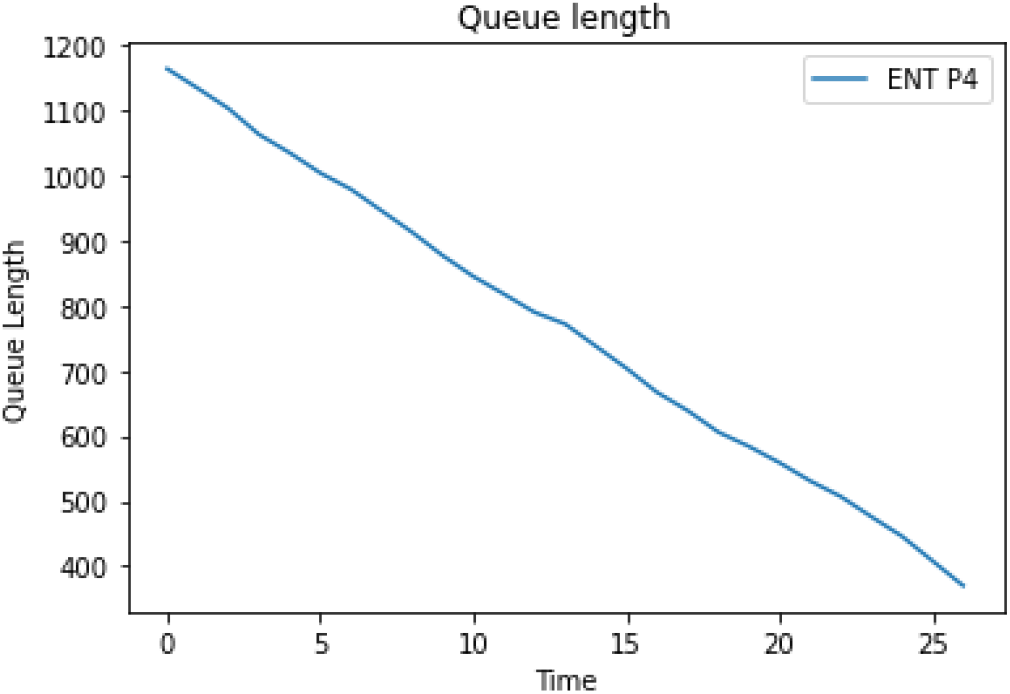
The reduction in waiting lists over the next 6 months under relief capacity is reduced to the target level of 390.

**Figure 4.**
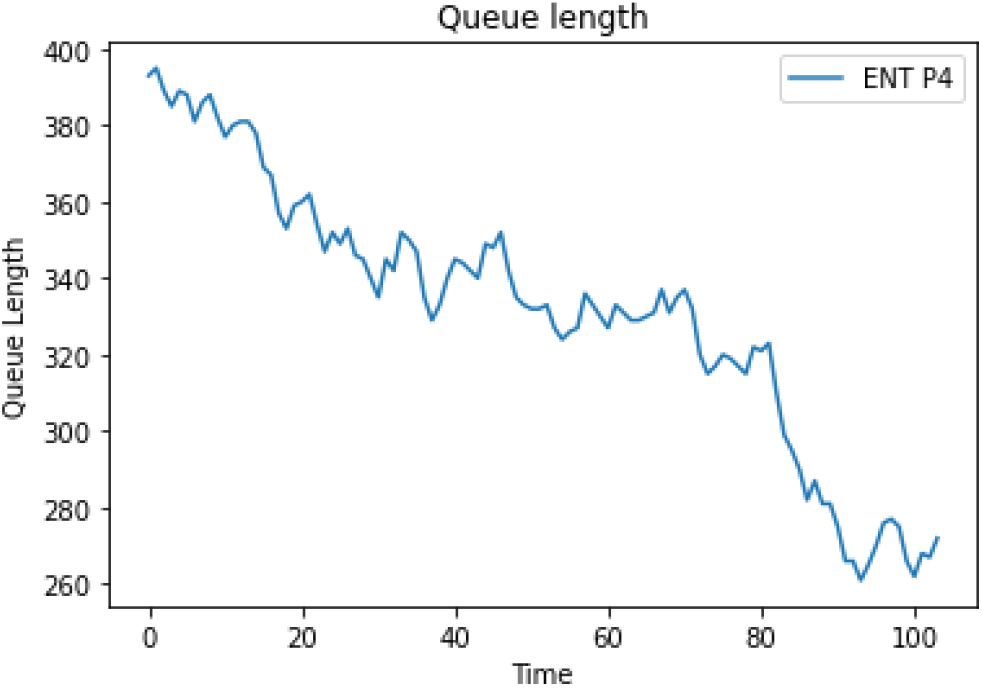
The queue process over the next year once the target equilibrium is reached. The mean wait of patients was 12 weeks and no patients missed targets.

A consequence is if we let 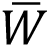 be the mean waiting time, then the probability of a given waiting time *W* exceeding this average is given by

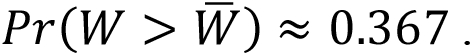

A property of exponential decay is that if we consider the chance that the waiting time is double the mean, then we square the above probability:

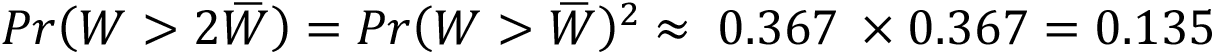

Continuing, similar reasoning holds if we triple or quadruple the chance of waiting above the mean. In particular, for four or six times the mean, we have:

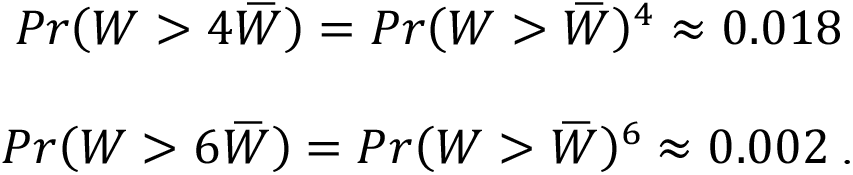

With this we can derive Fact 4 below: if we want to have a chance of less than 1.8% of missing the target, then the mean waiting time should be less than quarter of the target waiting time.

**Fact 4:** If we want to have a chance between 1.8%-0.2% of making a waiting time target, then the average patient should have a waiting time between a quarter and a sixth of the target. That is, we should aim to have:

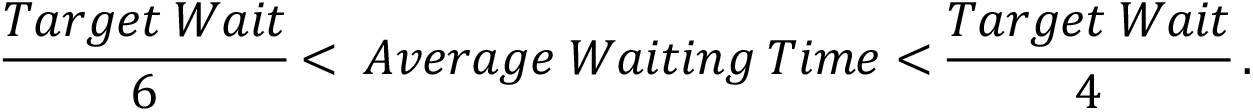

In the case of a P4 waiting list, the target wait is 52 weeks. Thus, we should expect the average patient being operated on to have waited between 9 and 13 weeks. In the case of P2 customers, the target is 4 weeks. Thus, the mean wait of a typical patient should be under one week.

Fact 4 gives a prudent rule for the target equilibrium that should be maintained across different waiting lists.

## Target Queue Length

While waiting times must be measured over time, the size of a waiting list can be measured instantly. A further indicator of the health of a waiting list is its queue size. A renowned result in queueing is Little’s Law which relates the queue size to waiting times through the demand for the waiting list (6)(7). Little’s Law requires relatively few assumptions, and, for this reason, it is one of the most famous and robust formulas in the field of operations research.

**Fact 5. [Little’s Law]** If Capacity > Demand then

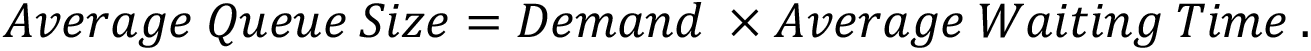

Thus if, as given in Fact 4 above, we want the average waiting time to be a quarter of the target, then Little’s Law gives us the following Target Queue Size.

**Fact 6. [Target Queue Size]**

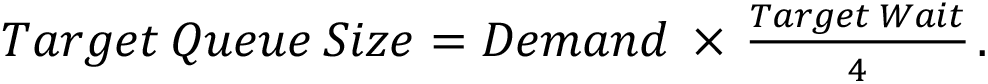

If the waiting list size is over twice the Target Queue Size, then we consider the queue to be sufficiently away from its target equilibrium that interventions are required. Therefore, we must increase capacity allocated to that waiting list before the queue sizes and waiting times get too large.

In our example, the demand is 30 patients a week and the target wait is 52 weeks. Thus, the target queue size is 30 × 52/4 = 390. So, in addition to not having enough capacity allocated, we can now see that the queue size of ENT P4 of 1200 is around three times bigger than it should be. Thus, special measures are required to reduce waiting times.

To summarize so far: we now know how big the queue size of a waiting list should be; we know how long the typical patient should wait; we know that the capacity must be larger than the demand on the waiting list. But how much larger should the capacity be? We consider this in two cases: the first, when the queue size is too big, specifically when the queue size is over double its target queue size. The other where the queue size is less than double its target.

## Relief Capacity

Whenever the queue size goes over twice the target queue size, the queue is at risk of missing its targets and a manager should direct capacity to that waiting list to reduce its size. We discuss how much capacity should be allocated.

Here we decide on a date by which the queue should be brought back to target levels, e.g. we may want to reduce the waiting list over 6 months and thus the target date 26 weeks from today’s date. Given the demand, we allocate sufficient capacity so that the target queue size is reached.

**Fact 7: [Relief Capacity]** If

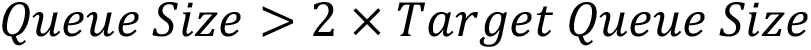

then decide on target date to relieve the queue and apply the capacity

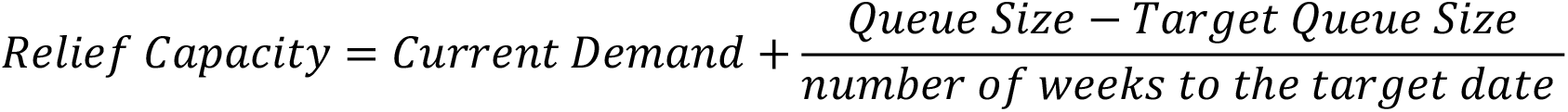

The relief capacity should be maintained until the target date or when the target queue size is reached.

The relief capacity will bring the queue down to target levels. In our example, the current queue size is 1200; the demand is 30 patients a week; the target queue size we found to be 390 and we decide a target of 26 weeks from today’s date (just before the beginning of the summer) to reduce the waiting list. Thus, the relief capacity is 30 + (1200-390)/26 = 61.15. So, we see that we require over double the number of ENT operations conducted to get the waiting list down to target levels over the next 6 months.

Although we consider the relief capacity to be a fixed amount of operation above the currently observed demand, we note that the relief capacity can be applied on a week-by-week basis. Here the demand is the number of referrals made the previous week and the relief capacity is demand plus the number of additional operations to be scheduled.

As discussed above if the queue size (or waiting time) is double its target mean then the capacity of the waiting list should be increased to reduce the queue to the target level. However, once the queue size is within an acceptable range, we can maintain the waiting time target with what is potentially a much smaller capacity allocation to the waiting list.

## Target Capacity

We have presented the waiting time and queue size of a waiting list operating at its target equilibrium. Now we discuss how many operations must be conducted each week to achieve the target equilibrium.

If the target queue size is achieved, then we can calculate a capacity allocation that will ensure waiting time targets in the long run.

**Fact 8: [Target Capacity Formula]** The target capacity is

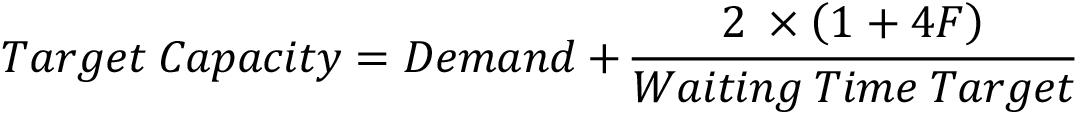

Here F is a parameter that depends on the variability in service. (If *F* is not known then it is usually reasonable to take *F* = 1.)

The above formula is a consequence of a well-known formula called the Pollaczek-Khinchine Formula (8).

A key observation is that if we add a constant amount of capacity above to the measured demand, then over the long run we can confidently make the waiting time targets. We will define *F* shortly, but for now we consider the case *F* = 1. In this case, 2 × (1 + 4*F*) = 10 in the above formula. For P2 waiting list priorities, the target wait is 4 weeks and so the target capacity is the weekly demand plus an additional 10/4=2.5 operations. That is, for any P2 waiting list, every four weeks there should be an additional 10 operations above the observed demand in those four weeks. For P3, the target capacity is the weekly demand plus 10/12. That is, for P3 waiting lists, every 12 weeks there should be an additional 10 operations above the demand in those 12 weeks. Similarly of P4, there should be 10 more operations than arrived over the past year. In the final case we begin to see that we need to distribute our capacity quite evenly to match the demand so that those 10 excess operations can take effect. To do this we must monitor the variability in capacity.

The term *F* can be calculated as follows: suppose *C* is the current number of operations per week; *V* is the current variance in the number of operations per week; *D* is the observed demand.

Define

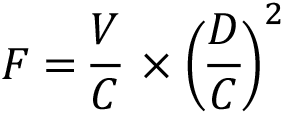

Here we see that the variance in the number of operations each week is an important factor in allocating sufficient capacity. For example, when we allocate 28 operations every 4 weeks, it is better to have 7 operations each week, than to do all 28 operations at the beginning of the month. Obviously, there is a trade-off here as it is often better for a surgeon to perform several operations over the course of a day or half day. However, beyond this it is worth keeping in mind that after capacity is fixed: **it is better to keep a well synchronized set of times to perform operations since this will have a beneficial impact in reducing waiting times**.

This can be measured through the value of F, which plays a similar role to the load. If F is less than 1 that is good. If F is greater than 1 then there is high variance in the capacity being allocated which may well result in large waiting times. A load less than 1 is essential to manage a waiting list, whereas F greater than 1 maybe acceptable (particularly when queue sizes are small) but could also be a symptom of problems in processing operations. Further, it is worth noting that (once the variability F is fixed) the amount of additional capacity required is a constant amount above the demand. In terms of staffing and theatre usage, this suggests that capacity should always be kept a constant amount above the observed demand. There will be instances where this capacity it not fully utilized, because the list is empty. But, as discussed, this availability is an important aspect of maintaining a healthy waiting list.

In the case of our ENT example, we see that the demand is *D* = 30 patients per week, capacity is *C* = 27 patients a week. The load is greater than 1and also the queue size is over twice the target, so the waiting list needs more capacity. The variance for operations each week (which is the square of the standard deviation) is *V* = 12 × 12 = 144. So

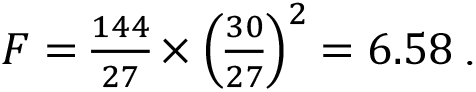

*F* is significantly greater than 1 which is a symptom of both overloading and high variability in the number of operations. The target wait is 52 weeks, so we deduce that the ENT capacity should be at least

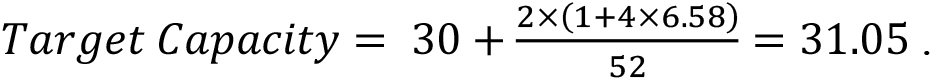

Here, we see that if each week we do roughly 1 more operation than arrives then the waiting target will be met. The above target capacity is a relatively cautious bound.

## Waiting Pressure

Finally, what are the trade-offs between different waiting lists and their targets? For instance, there may not be sufficient capacity for a set of waiting list. Then which waiting list should be prioritized to best make use of the capacity that is available? Or conversely, suppose that there is unused capacity because a waiting list is empty. To which waiting list should that free capacity be allocated?

A metric that can be used is the pressure, which is essentially the proportion of the target that has elapsed on a given waiting list.

**Fact 9 [Waiting List Pressure]**. For a waiting list with target waiting time, the pressure on the waiting list is twice the mean delay divided by the waiting list target

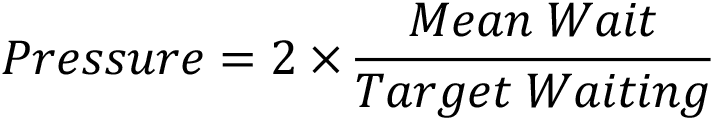

The pressure of any given waiting list should be less than 1. If the pressure is greater than 1 then the waiting list is most likely going to miss its target.

The pressure effectively gives a “currency” by which we can compare waiting list targets. For instance, let’s compare the ENT P4 waiting list with the ENT P2 waiting list. The ENT P2 waiting list has a target of 4 weeks and there are currently 220 patients waiting for their operation. The average delay of a patient is 24 weeks. Whereas there are 1200 ENT P4 patients which have a target of 52 weeks with an average delay of 119 weeks. The pressure on ENT P2 procedures is 24/4 = 6 whereas the pressure on ENT P4 procedures is 61/52 = 1.17. We see that both lists have a pressure over 1 and thus are missing targets, we see that ENT P2 is significantly higher than P4 procedures thus a closer attention should be paid to P2 procedures. (The capacity and target queue sizes can then be calculated as above.) We can use the pressure rank to compare lists further. For instance, in the case of ENT P2, the pressure can be calculated for different hospitals. The pressure for ENT P2 at Hospital A is 17 whereas the pressure for ENT P2 at Hospital B is 3. This suggests that where possible new procedures should be referred to Hospital A to balance the pressure between the two sites.

We have now summarized several key metrics that can be used to understand and improve the efficiency of waiting lists. We now develop this further with several examples.

## Case studies

We provide some representative data of how the above formulas and metrics can be used to understand waiting list pressures at different levels for procedures, for specialties, for hospitals or for hospital trusts.

### Analysis of Specialties at Hospitals

Next we consider the allocation of three specialties which we label S,T,U. We then consider the allocation of one of these specialties among three hospitals which we label M,N,O.

A table is given in Figure 6. This gathers the required queueing metrics that the trust can then use to understand waiting lists and their requirements. This example is a representative sample of a much larger table that is used for making capacity planning and routing decisions for waiting lists.

**Figure 6.**
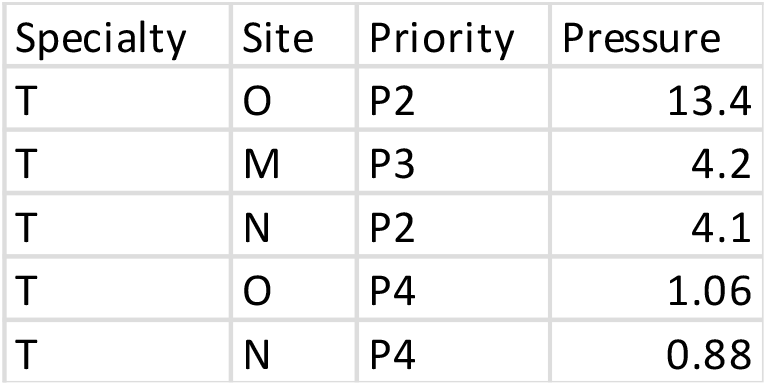

**Figure 9.**
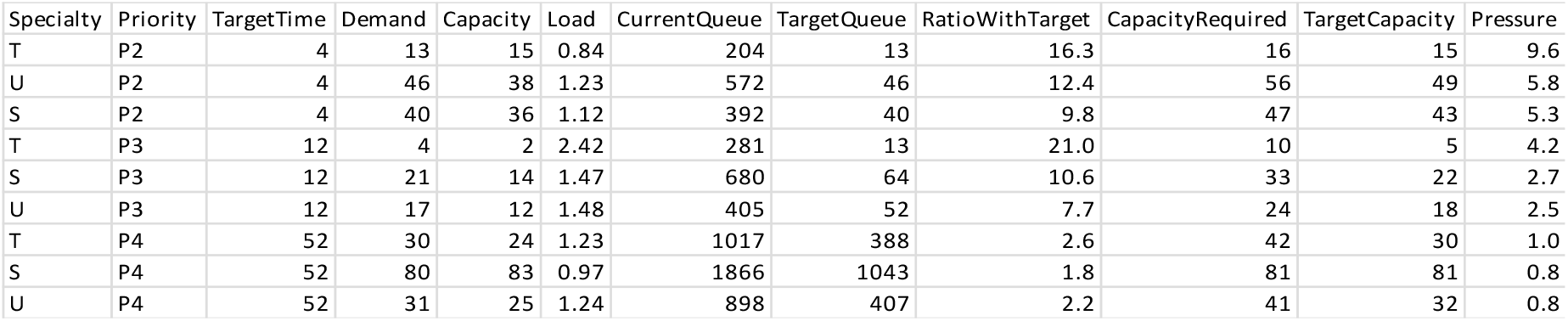

Here the waiting list considers the three specialties with each priority group. It is ordered by the pressure on these services. We see that Specialty T consistently has the highest pressure. On two entries, Specialty S Priority 4 and Specialty U Priority 4 have a pressure below 1, which is a requirement to meet targets. Only Specialty S Priority 4 is in good condition having a load less than 1 and a queue size within a ratio of 2 of the Target Queue Size. This is the only waiting list that does not require some form of intervention. The capacity requirements for these queues are given. When the queue size ratio is above 2, this recommendation is the rate at which operations need to be conducted to reduce waiting lists to target levels within a year otherwise it is the Target Capacity needed to maintain targets with each queue in equilibrium.

Focusing on Specialty T which has the highest waiting list pressure, we can look at the pressure for this specialty at different hospitals. Here in Figure 6, we see that the waiting lists at Hospital O are under higher pressure than Hospital N. This suggests that Specialty T procedures should be moved from Hospital O to Hospital N wherever possible.

In general, the waiting list pressures across different sites should be balanced to ensure resources are pooled together to achieve a consistent level of service. Capacity should be attributed as above so the pressures are less than 1.

### Analysis of Procedures

The Hospital N has a new operation model designed by managers for non-covid elective operations. This has increased the capacity of Hospital N and it can now provide a range of procedures that it could not previously. Rather than consider how much capacity needs to be allocated, as we did above, the question now is how this newfound capacity should be assigned given the current waiting lists?

The capacity planning calculations discussed above are applicable for specialties where there are tens to hundreds of referrals each week. However, many procedures are highly specialized often with no referrals in each week. A simple way to reroute procedures is to rank the waiting list pressure for different procedures and then assign the highest ranked operations. Again, this will have the effect of equilibrating the pressures across different procedures.

Here we analyzed 250 procedures, Operation Groups in Specialties S, T and O. Ranking them from highest pressure to lowest. The average pressure is 4.1 with 225 procedures having a pressure above 1, suggesting that without intervention these waiting lists will not meet their targets for most patients. Given the available capacity the highest ranked procedures are recommended to be rerouted to the new facility at Hospital N.

## Other Considerations

So far, we have focused on constructing metrics, targets, capacity allocations and scheduling decisions. We now discuss other factors that go beyond the concepts introduced so far. Each of these can be modelled, analyzed, and optimized to improve efficiency. We detail some aspects of these decision-making processes.

### Staffing

The staff levels required can be analyzed in a similar way that we model the demand and capacity for a queue. There is a demand for staff for operations, which the demand for each operation multiplied by the number of staff each operation requires. There is a capacity for each group of staff. That is the number of operations that they can perform each week. We can then plan the amount of capacity required to meet this demand and achieve low queue sizes. Such a system requires quite detailed information on the requirements for each procedure and for this reason we do not investigate that here. However, we note that the Waiting List Pressure can be used for skill-based routing of patients to consultants (9).

### Theatre Scheduling

Here we do not schedule operations but instead determine the number of operations required to make targets. It is possible to analyze theatre occupancies and then schedule this for optimal efficiency (10). Surges using overtime can be used to help improve scheduling (11). However, there are specialist theatres within hospital trusts that are in almost continuous use. For elective operations the current bottleneck is staffing levels. There are numerous factors which provide a source of inefficiency in scheduling operations, such as the mismatch between the scheduled time of an operation and the time actually taken, which, although clinically necessary, can lead to some predictable inefficiency when scheduling.

### Routing

We have discussed routing of patients using waiting list pressure. This is a robust rule used in many queueing systems (12). However, more static optimization can be considered where proportions of demand for different specialties can be referred across a trust to improve efficiency.

### Demand estimation

We have used a relatively simple estimate of demand for services based on recent demands over the last 3 months. However, more advanced regression methods can be used to improve demand estimates and thus consequently improve the predictive power of formulas applied here (13). With more powerful estimation of demand, there is greater potential to plan and optimize the utilization of hospital resources.

### Queueing Discipline

Certain metrics can be improved through different disciplines applied within a queue. For instance, prioritizing short operations ahead of long operations can reduce the number of people waiting in a queue (14). However, here there is a trade-off as the waiting time for those left in the queue will increase considerably. For reasons of fairness, we focus efforts on creating systems that are close to a first-come first-served discipline within each priority group.

## Conclusions

In summary, this paper introduces queueing theory for managers and clinicians. We describe the basic mathematical concepts that govern the behaviour of NHS waiting lists. The implications of queueing theory are often less than intuitive, and sometimes even counterintuitive, but recognising this is an important element in recognising the scale of the problem and the scale of resource required to address it.

The use of targets for waiting lists for NHS lists is appropriate. However, there must be mechanisms in place to understand how to achieve these targets. To this end, we would advocate the need for national standards for assessing and managing waiting lists. By analogy, medical trials require strict standards of case and must be demonstrably effective with a high level of statistical precision. Given the equal importance of waiting times on health outcomes, it is a reasonable aim that waiting lists are managed with comparably rigorous standards.

## Data Availability

All data produced in the present work are contained in the manuscript.

